# Turns and Downturns in Aging Drivers

**DOI:** 10.64898/2026.02.04.26345564

**Authors:** Marie Hardt, Guillermo Basulto-Elias, Heike Hofmann, Shauna Hallmark, Anuj Sharma, Jeffrey D. Dawson, Matthew Rizzo, Jun Ha Chang

**Affiliations:** Department of Statistics, Iowa State University, 2438 Osborn Dr., Ames IA 50011, USA; Institute for Transportation, Iowa State University, 2711 S. Loop Drive Suite 4700, Ames IA 50010, USA; Department of Statistics, University of Nebraska-Lincoln, 340 Hardin Hall North Wing, Lincoln NE 68583, USA; Department of Biostatistics, University of Iowa, 145 N. Riverside Dr., Iowa City IA 52242, USA; Department of Neurological Sciences, University of Nebraska Medical Center, 4242 Farnam St. Suite 650, Omaha NE 68131, USA

**Keywords:** Older drivers, Cognitive decline, Naturalistic driving study, Left-turning behavior, Digital behavioral markers, Mild cognitive impairment, Alzheimer’s disease

## Abstract

As cognitive decline progresses, older adults may self-regulate their driving. Avoidance of left turns across traffic is observable in naturalistic driving data but rarely self-reported. We studied 106 older adults using baseline and one-year follow-up neuropsychological assessments. In-vehicle sensors passively recorded driving behavior over 12 weeks. We identified 295,112 turns from vehicle heading changes. We used mixed-effects logistic regression to model the odds of turning left, with cognitive status category change from baseline to one-year follow-up as the predictor. Greater cognitive impairment, represented by movement to a more severe cognitive status category at one-year follow-up, was associated with reduced odds of turning left (odds ratio = 0.984, 95% confidence interval = 0.969–0.999; P value = .037). Left-turn avoidance may be a behavioral marker of early cognitive decline. Passive driving data could help detect functional changes, enabling intervention to preserve mobility and independence. Further research is needed to establish a clinical threshold of concern for decreasing trends in left turn frequency in older drivers.

## 1. Background

Early detection of cognitive decline, particularly mild cognitive impairment (MCI) and Alzheimer’s disease (AD), is a critical priority for clinical care and public health. While neuropsychological testing remains the gold standard, there is growing interest in identifying real-world behavioral biomarkers that reflect subtle, prodromal changes. Driving—a cognitively demanding activity involving attention, visuospatial processing, executive function, and motor coordination—offers a natural context to observe such changes.

Naturalistic driving data allow for the passive capture of decision-making and motorist behavior in everyday settings. In a review of the literature available as of 2022, Bayat and Roe noted that several naturalistic driving studies involving older drivers, where data was collected by the Driving Real-world In-Vehicle Evaluation System (DRIVES), showed that older drivers with preclinical AD drove less often than their cognitively normal counterparts and were also less likely to adopt aggressive driving behaviors like hard braking, speeding, and sudden acceleration (Bayat and Roe 2022). Intersections, in particular, pose significant challenges for older drivers due to their high cognitive and perceptual demands. Age-related cognitive decline increases crash risk, especially during complex maneuvers like left turns (Basulto-Elias et al. 2023).

Older drivers are disproportionately involved in intersection crashes. For example, those aged 65–69 are 2.3 times more likely and those aged 85+ are 10.6 times more likely than younger drivers to be involved in a fatal intersection crash (Preusser et al. 1998). Naturalistic driving studies such as that conducted under the Second Strategic Highway Research Program (SHRP 2) have confirmed an elevated intersection crash risk in drivers over 70 (Seacrist et al. 2020). The risk continues to rise with age and is further amplified in drivers with cognitive disorders. A recent meta-analysis identified intersection navigation as one of the most common sources of errors in drivers with dementia (Camilleri and Whitehead 2023).

Left turns are particularly hazardous. Older drivers are frequently cited for failure to yield, especially at stop-controlled intersections, where older drivers are significantly more likely than younger drivers to be at fault in a crash (Sifrit et al. 2011; Braitman et al. 2007). Failure-to-yield errors often occur during left turns and are more common among drivers 70 and older (FHWA 1995).

Some older drivers compensate for perceived deficits by avoiding nighttime driving or high-traffic environments. However, avoidance of left turns, a cognitively and perceptually demanding maneuver, is rarely self-reported (Molnar and Eby 2008; Molnar et al. 2013; Beck, Luo, and West 2022), despite being observable in driving data.

A recent study from the Longitudinal Research on Aging Drivers (LongROAD) project showed that the monthly ratio of right turns to left turns was among the most predictive features in classifying drivers with MCI or dementia (Di et al. 2023). This suggests that passive, sensor-based monitoring of turning behavior can capture subtle behavioral changes not reflected in self-reporting.

While the LongROAD study provided important insights, it relied on cross-sectional classification based on the most severe cognitive state and excluded participants with shorter study participation. It also did not distinguish between MCI, dementia, and AD.

Because driving involves complex, goal-directed behavior, early-stage cognitive dysfunction may manifest as changes in turning behavior long before a formal diagnosis is made. Identifying these changes could support the development of behavioral biomarkers for cognitive impairment.

Additionally, understanding the driving challenges faced by older adults can inform roadway design and traffic control strategies to enhance safety.

This study evaluates whether older drivers with cognitive decline exhibit reduced odds of making left turns. By comparing turning behaviors among individuals with and without cognitive decline, we assess the potential use of left-turn frequency as a real-world marker of early cognitive change.

## 2. Methods

### 2.1 Study Design and Data Collection

This study used interim data from an ongoing naturalistic driving study sponsored by the University of Nebraska Medical Center (UNMC). The parent study was designed to include a baseline cognitive assessment and the collection of three months of real-world driving behavior using in-vehicle sensors and wearable devices, followed by annual cognitive assessments over a two-year period. At the time of this analysis, data from the baseline and one-year follow-up assessments were available.

Each participant’s vehicle was equipped with a custom-built sensor system (a vehicle Black Box system) that passively recorded data from when the vehicle ignition was turned on to when it was turned off. Each ignition cycle (on to off) defined a single “drive.” The system included forward-facing roadway and cabin cameras, a Global Positioning System (GPS) unit, and an inertial measurement unit (IMU). On-board diagnostic (OBD) data, such as throttle position and engine revolutions per minute (RPM) were also recorded. Data were sampled at 1 Hz (i.e., one observation per second). The UNMC Institutional Review Board approved the protocol (0522-20-FB), and informed consent was obtained from all participants.

We note that standardized road tests and driving simulators are frequently used to assess driver performance. However, these data sources utilize identical real-world driving courses or virtual driving scenarios. We would then expect that each study participant would complete the same number of left and right turns as they navigated the course, whether in the real world or in a simulator. Since these driving scenarios are controlled and practically the same for each study participant, there would be no value in analyzing the number of left and right turns taken by the participants in these scenarios. In contrast, naturalistic driving studies permit participants to drive as they normally would. The drivers make choices about their routes (whether consciously or unconsciously) and thus make an analysis of turning behavior meaningful.

### 2.2 Participants

We enrolled 182 active, legally licensed drivers aged 65 to 90 years from the Omaha, Nebraska area. Participants were recruited through social media, newspapers, flyers, and outreach to community organizations (Merickel et al. 2019).

All participants met Nebraska’s visual acuity standards (>20/50 OU, corrected or uncorrected) and self-reported no exclusionary medical conditions (e.g., dementia, heart failure, psychiatric illness) or use of psychoactive medications. Common age-related conditions like arthritis were not exclusionary. Participants completed baseline and one-year follow-up questionnaires on demographics, driving experience, medical history, and socioeconomic status. Table 1 summarizes the participants’ characteristics and the turning endpoints.

**Table 1:** Demographic characteristics and turning endpoints, overall and by cognitive prognosis class.

| Characteristic | Value | Overall,<br>n = 106 | Normal<br>→<br>Normal,<br>n = 43 | Normal<br>→ MCI,<br>n = 16 | MCI →<br>MCI, n<br>= 36 | MCI →<br>AD, n =<br>11 |
| --- | --- | --- | --- | --- | --- | --- |
| Age, mean (SD),<br>years |  | 75.3 (6) | 74.1<br>(5.1) | 74.2 (6) | 76.9<br>(6.7) | 76 (5.9) |
| Sex | Female | 61<br>(57.5%) | 33<br>(76.7%) | 8 (50%) | 16<br>(44.4%) | 4<br>(36.4%) |
|  | Male | 45<br>(42.5%) | 10<br>(23.3%) | 8 (50%) | 20<br>(55.6%) | 7<br>(63.6%) |
| Race | White | 102<br>(96.2%) | 43<br>(100%) | 16<br>(100%) | 33<br>(91.7%) | 10<br>(90.9%) |
|  | Black | 3 (2.8%) | 0 (0%) | 0 (0%) | 3 (8.3%) | 0 (0%) |
|  | American<br>Indian | 1 (0.9%) | 0 (0%) | 0 (0%) | 0 (0%) | 1 (9.1%) |
|  | Asian | 0 (0%) | 0 (0%) | 0 (0%) | 0 (0%) | 0 (0%) |
| Employment | Retired | 82<br>(77.4%) | 30<br>(69.8%) | 11<br>(68.8%) | 33<br>(91.7%) | 8<br>(72.7%) |
|  | Employed full/part time | 15<br>(14.2%) | 8<br>(18.6%) | 4 (25%) | 2 (5.6%) | 1 (9.1%) |
|  | Volunteer | 8 (7.5%) | 5<br>(11.6%) | 0 (0%) | 1 (2.8%) | 2<br>(18.2%) |
|  | Keeping house | 1 (0.9%) | 0 (0%) | 1 (6.2%) | 0 (0%) | 0 (0%) |
| Driving Experience, mean (SD), years |  | 58.6<br>(6.6) | 57.9<br>(6.6) | 56.4<br>(6.3) | 60.1<br>(6.8) | 59.4<br>(6.3) |
| Weekly Driving, mean (SD) | Miles | 85.3<br>(75.4) | 84.7<br>(78.6) | 91.1<br>(91.3) | 81 (71.1) | 93.6<br>(57.8) |
|  | Kilometers | 137.2<br>(121.3) | 136.3<br>(126.5) | 146.6<br>(146.9) | 130.3<br>(114.4) | 150.6<br>(93) |
| Number of Turns Detected Per Participant, mean (SD) | Left | 1394.6<br>(664.9) | 1330.3<br>(598.3) | 1616<br>(857.7) | 1414<br>(649.2) | 1260.3<br>(665.2) |
|  | Right | 1389.5<br>(650) | 1303.2<br>(562.9) | 1602.5<br>(886.2) | 1421.3<br>(618.4) | 1313.1<br>(689.1) |
| Percentage of Turns Detected Per Participant, mean (SD) | Left | 49.9%<br>(2.2%) | 50.3%<br>(1.8%) | 50.3%<br>(1.7%) | 49.6%<br>(2.6%) | 49%<br>(2.8%) |
|  | Right | 50.1%<br>(2.2%) | 49.7%<br>(1.8%) | 49.7%<br>(1.7%) | 50.4%<br>(2.6%) | 51%<br>(2.8%) |

NOTE. Percentages may not add to 100% due to rounding.

### 2.3 Measures

#### 2.3.1 Turn Detection

We identified turning behavior using gyroscopic heading data, which are more reliable than GPS data at the low speeds typical of intersections. A turn was defined as a change in heading exceeding ±10^°^ within one second. More specifically, let *h*_*t*_ *be a t*ime series of directional measurements (i.e. *h*(*t*) ∈ (−180^°^, +180^°^)), observed at discrete time points *t*_*i*_ with *i* ∈ *N* spaced at one second intervals. A turning event is then identified when a change in heading *h*′(*t*): = *h*(*t* + 1) − *h*(*t*) exceeds the bounds of an interval [λ, ρ], where λ and ρ, with λ ≤ ρ, are the most extreme angle values (in degrees) still considered to indicate driving on a straight path. In other words, any movement sufficiently different from maintaining a straight path was detected as a turning event. To avoid overcounting during continuous turning, we counted only the first instance in sequences of large heading changes.

Figure 1 illustrates our turn detection criteria. All turning maneuvers, regardless of location (e.g., intersections, parking lots), were included. Assuming that a car is heading eastbound at time *t*_0_with heading *h*(*t*_0_) = 0, the shaded cone in Figure 1 represents the allowable potential locations (*x*1, *y*1)that can be reached while the vehicle is driving on a straight path 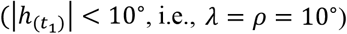 at a speed of up to 50 mph. Movements outside the cone representing a straight path are classified as turning events. Vertical arrows depict the lateral offset from the straight path at different speeds. For example, at 30 mph, a heading deviation of 10^°^ results in more than half a lane shift within one second. Corresponding numerical values for the lateral offsets are presented in Table A.1 in the appendix.

**Figure 1:**
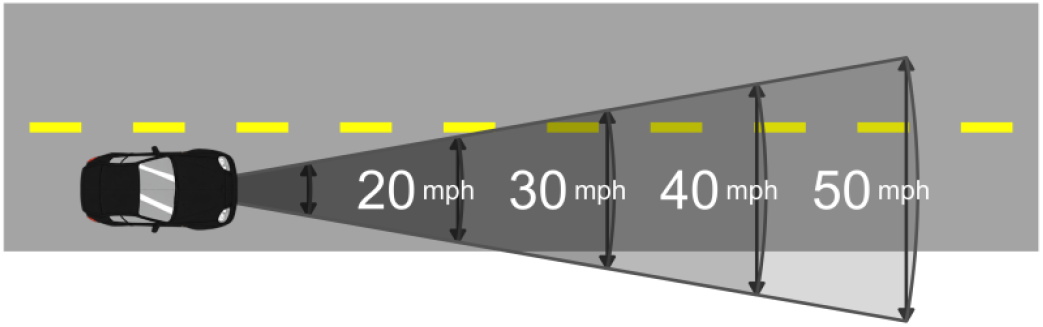
Turn detection criteria at various speeds based on heading deviation.

Figure 2 and Figure A.1 (in the appendix) depict left and right turn detection in different driving contexts. Each dot represents the GPS location of the vehicle at one-second intervals. The blue dots highlight the locations at which the change in heading went beyond the −10^°^ threshold for a left turn and thus left turns were detected. Similarly, the orange dots highlight the locations at which the change in heading exceeded the +10^°^threshold for a right turn, thus resulting in a detected right turn. Note that the GPS locations do not always align precisely with the road geometry, which further motivated our use of gyroscopic heading direction instead of GPS location to identify turns. Although our turn detection method is simple, the large numbers of detected left and right turning maneuvers over the three-month driving data collection period give a stable estimate of each participant’s driving behavior.

**Figure 2:**
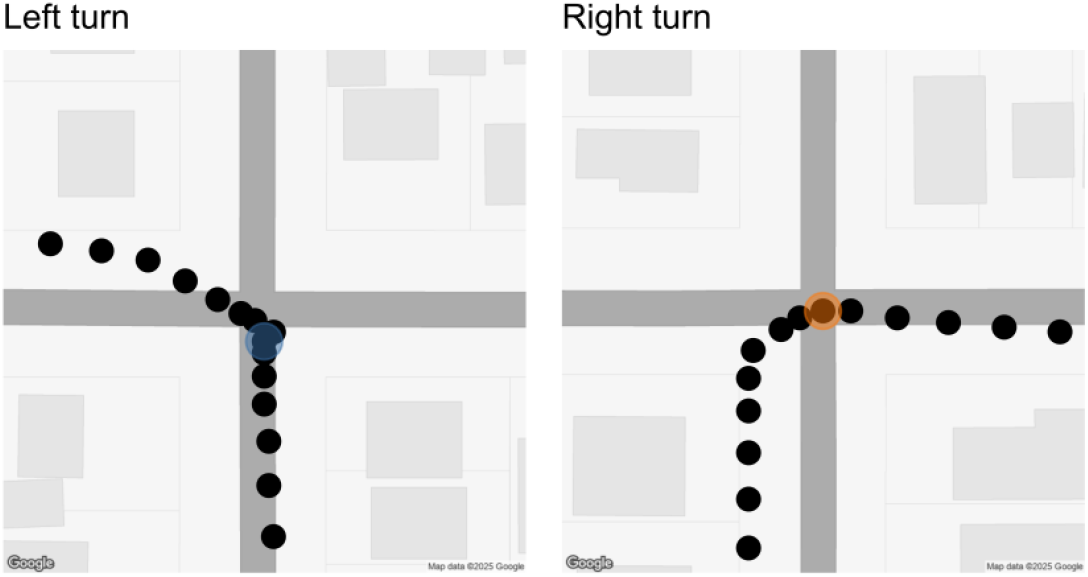
Left (blue) and right (orange) turns detected during a northbound drive.

#### 2.3.2 Cognitive Status Classification

Cognitive status was assessed through a baseline assessment and a one-year follow-up assessment using standardized neurological and neuropsychological evaluations (Albert et al. 2011), which included domains such as memory, language, visuospatial skills, executive function, and attention. Classification into categories for normal cognition, MCI, or dementia was based on National Institute on Aging - Alzheimer’s Association (NIA-AA) and National Alzheimer’s Coordinating Center (NACC) Uniform Data Set (UDS) version 3 criteria, including the Petersen/Winblad and Jak/Bondi definitions (Petersen et al. 1999; Jak et al. 2009).

Functional assessments used the Clinical Dementia Rating (CDR) and Functional Activities Questionnaire (FAQ) (Morris 1993; Marshall 2015). An FAQ > 9 and a CDR ≥ 0.5 indicated functional decline.

Several criteria were used to exclude participants from the model. We excluded N=50 participants from the model because they had not yet completed the one-year follow-up cognitive assessment. Participants were also excluded if they showed cognitive reversion (N=14) at the one-year follow-up assessment or dementia at the baseline assessment (N=7), which focused the analysis on normal and MCI trajectories. As shown in Figure 3, the majority of participants who did not exhibit cognitive reversion had a period of up to 15 months between the end of the driving data collection period and the one-year follow-up cognitive assessment. To ensure similarity in the amount of time from the end of the driving data collection period to the one-year follow-up cognitive assessment, N=4 participants were excluded because they had more than 15 months from the end of driving data collection to the one-year follow-up assessment. Another participant who had only 61 days between the end of the driving period and the one-year follow-up assessment was also excluded because these dates were much closer than for any other participant. This participant is not shown in Figure 3. A total of 106 participants were included in the model. Final classifications included four cognitive trajectory groups based on baseline and follow-up status:

**Figure 3:**
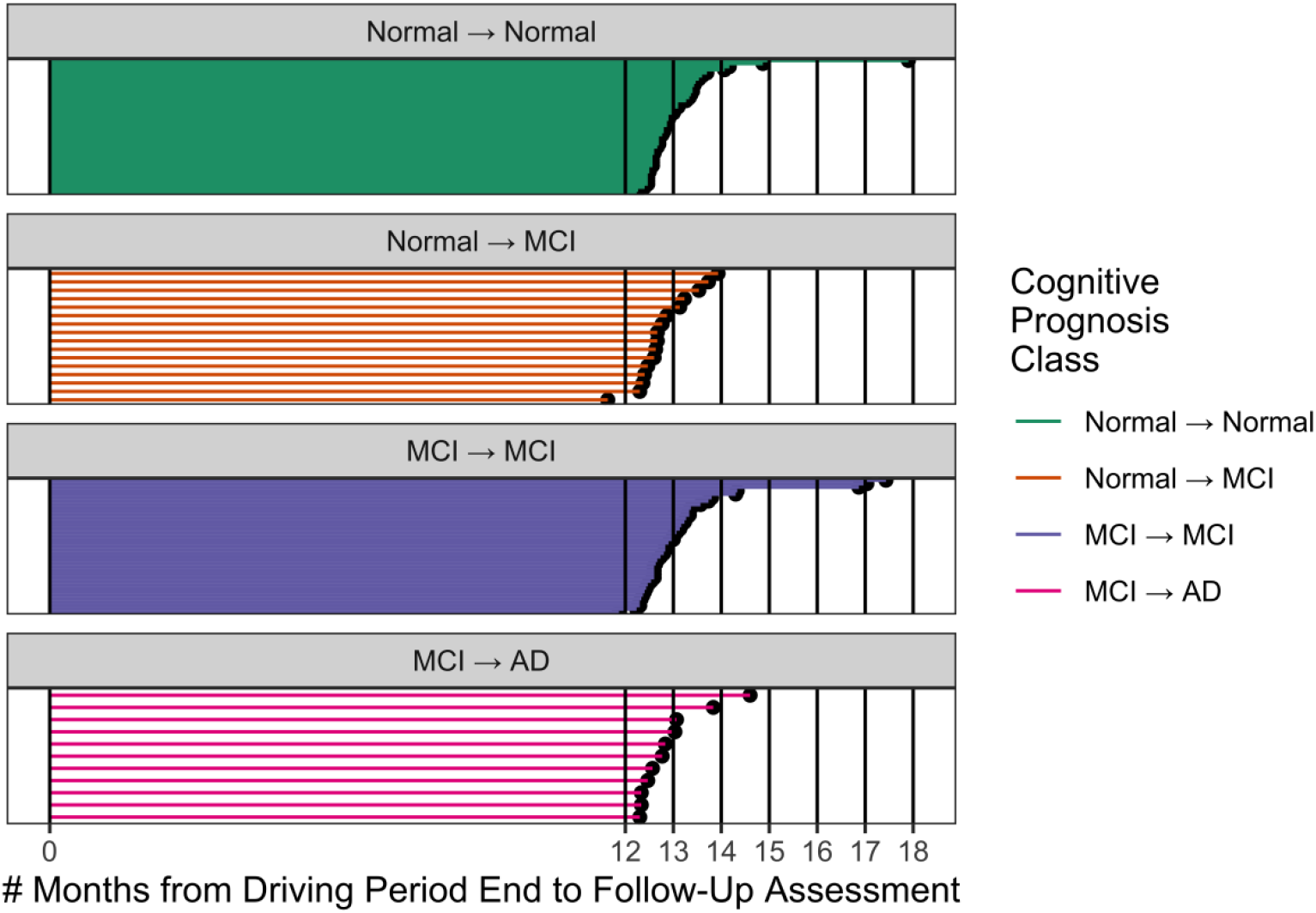
The number of months between the end of the driving data collection period and the follow-up cognitive assessment was used to determine a cutoff of 15 months for inclusion in the model. This cutoff excluded 4 participants from the model.

1. Normal → Normal (stable normal)
2. Normal → MCI (incident MCI)
3. MCI → MCI (stable MCI)
4. MCI → AD (progression to dementia)

## 3. Statistical Analyses

To evaluate the relationship between turning behavior and cognitive prognosis, we used a mixed-effects logistic regression model, discussed in Section 3.1.

### 3.1 Mixed-Effects Logistic Regression Model

#### 3.1.1 Outcome and Predictor Variables for the Logistic Regression Model

The dependent variable for the logistic regression model was a binary indicator of turn direction: 1 for left turns, 0 for right turns. The independent variable was the cognitive prognosis class (1 through 4), treated as a numeric ordinal variable reflecting increasing severity of cognitive decline.

#### 3.1.2 Logistic Regression Model Specification and Justification

We used mixed-effects logistic regression to model the odds of a driver making a left versus a right turn, with participant-level random intercepts to account for repeated measures. We specify the Turn Model (TM1) as:

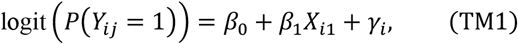

where *Y*_*ij*_, denotes the turn outcome (1 = left, 0 = right) for detected turn *j* made by study participant *i*, β_0_is a fixed intercept, *X*_*i1*_is the cognitive prognosis for study participant *i*, *β*_1_ is a fixed effect associated with the cognitive prognosis, and γ_*i*_ is a participant-level random effect. We assume that the random effect γ_i_ is normally distributed with mean 0 and variance 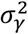. Note that, in this model, each turn is included as a separate event. This means that there is an implicit weighting for more active drivers. Even with a small overall sample size and small cognitive prognosis class sizes, the overall average of approximately 2800 left and right turns (combined) detected per study participant provides a large number of observations and lends power to the model.

We tested the assumption that cognitive prognosis could be treated as a numeric ordinal variable (rather than a categorical variable) using a likelihood ratio test (Williams 2020). The test was performed using the R function anova. The test statistic, χ^2^ = 0.457, df = 2, *P* value = .796, supported the simpler ordinal model.

Random intercepts accounted for between-subject variation; thus, we did not adjust for age, sex, or driving experience, but rather assumed that these were captured by subject-level variation. The model was fit in R using the glmer function from the package lmerTest (Kuznetsova, Brockhoff, and Christensen 2017).

#### 3.1.3 Logistic Regression Model Results

##### 3.1.3.1 Participants

Among the 106 study participants included in the model, our algorithm identified 295112 turns: 147825 were classified as left turns and 147287 as right turns. Participant demographic characteristics and turning endpoints are presented in Table 1.

Figure 4 shows boxplots of the percentage of left turns by cognitive prognosis group. The orange intervals represent 95% bootstrap confidence intervals for the mean (Efron and Tibshirani 1993). A decreasing trend in the percentage of left turns is observed with increasing severity of cognitive impairment. This is clearly seen in the median percentage of left turns for each cognitive prognosis class, denoted by the black vertical line inside each box. Variability also differs across the four cognitive prognosis classes, as indicated by the different lengths of the boxes and the whiskers extending out from the boxes for each cognitive prognosis class. The boxplots clearly show a relationship between cognitive prognosis class and turning frequency.

**Figure 4:**
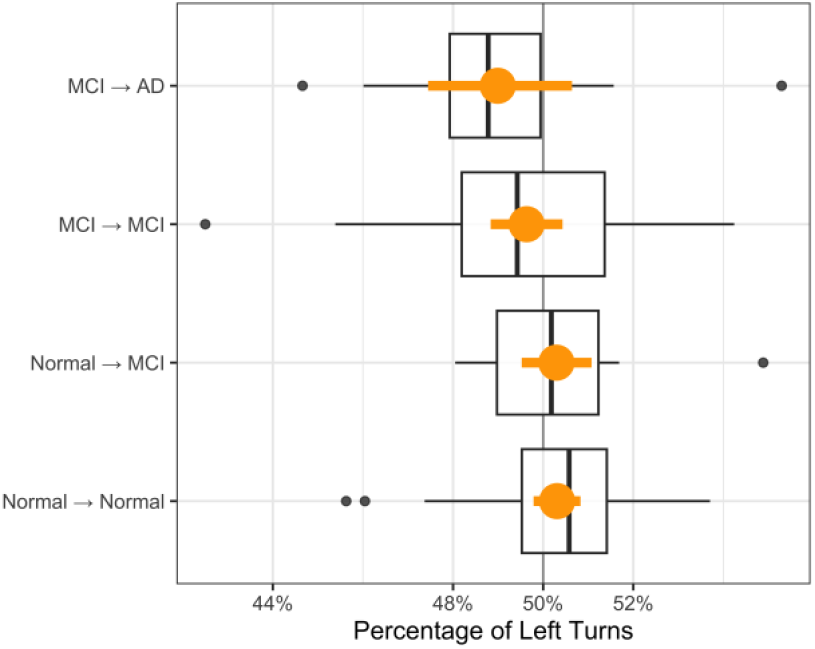
Boxplots of the percentage of left turns for each cognitive prognosis class with a cutoff of 15 months (450 days) from the end of the driving data collection period to the one-year follow-up assessment. The orange intervals are 95% bootstrap confidence intervals.

##### 3.1.3.2 Mixed-Effects Logistic Regression Model Estimates

The estimate of *β*_1_ (the fixed effect associated with cognitive prognosis class) is -0.016 with a standard error of 0.008. This leads to an odds ratio estimate of 0.984 with a 95% confidence interval of (0.969, 0.999) and a P value of .037. Thus, each unit increase in cognitive prognosis class (indicating progression to more severe cognitive impairment) is associated with a 0.984-fold decrease in the predicted odds of a driver making a left turn. Equivalently, such an increase in cognitive prognosis class is associated with a 1.016-fold increase in the predicted odds of a driver making a right turn. The estimate of 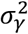 (the variance of the participant-level random effect) is 0.006. The model results suggest that as cognitive impairment progresses over time, older drivers become less likely to make left turns.

## 4. Discussion

### 4.1 Conclusions

This study investigated the relationship between cognitive status and the likelihood of making left turns among older drivers. Using naturalistic driving data and longitudinal cognitive assessments, we found that greater cognitive impairment was significantly associated with reduced odds a driver making a left turn. Conversely, a lower frequency of left turns was associated with greater cognitive decline.

Unlike prior research relying on self-reported avoidance of certain driving situations (e.g., (Molnar and Eby 2008; Molnar et al. 2013; Beck, Luo, and West 2022)), our approach utilized passively collected real-world driving behavior. Consistent with Di et al., who identified the ratio of right turns to left turns as a predictive feature of cognitive impairment, our findings reinforce the value of turning behavior as a digital behavioral marker (Di et al. 2023). Our longitudinal model strengthens this evidence by linking turn frequency to clinically verified cognitive trajectories. Importantly, our findings are based on changes observed over just one year. A longer-term longitudinal study using a second annual follow-up assessment is underway to determine whether these trends persist or evolve over time.

Our findings suggest a direction for future research. Future naturalistic driving studies involving older drivers can incorporate a larger study cohort, a longer driving data collection period, and a longer follow-up period. Detecting and assessing turning maneuvers with more sophisticated tools and methods will lead to a more comprehensive understanding of the relationship between turning behaviors and cognitive status in older drivers. This, in turn, will establish a threshold for clinical concern related to a decreasing trend in the frequency of left turns.

### 4.2 Limitations

This study has several limitations. First, the study cohort consisted of primarily white, community-dwelling drivers aged 65 to 90 from the Omaha, Nebraska area. This may restrict the generalizability of the analysis results to other older drivers of different demographics living in other areas.

Second, our heading-based method for detecting turns was simple and broad and may have included non-intersection maneuvers, such as navigating road curves or lane changes. Future work should explore more precise turn detection approaches. However, the large number of turns detected per study participant gives a stable estimate of each participant’s turning behavior throughout the driving data collection period.

Third, while we observed a relationship between cognitive prognosis and turning behavior, the effect was subtle. Babulal et al. noted that large sample sizes (e.g., 150 per group) may be necessary to detect meaningful behavioral differences between older adults with and without preclinical AD (Babulal et al. 2019). Our analysis included a total of 106 participants, which may have limited the significance of the findings. However, the large number of turns detected per participant provided a large number of observations used in the model.

Fourth, our mixed-effects logistic regression model considered only cognitive status and did not include other potentially important covariates, such as age, sex, or driving experience. The inclusion of the participant-level random effect in the model helped to mitigate any potential effect of other covariates. However, future studies should explore more comprehensive models that incorporate multiple covariates and utilize more advanced methods, such as random forests, to enhance predictive accuracy (Rizzo and Dawson 2025).

## Acknowledgements

We extend our sincere gratitude to the Mind & Brain Health Labs in the Department of Neurological Sciences at the University of Nebraska Medical Center for their leadership in recruiting participants for this study.

## CRediT Author Contribution Statement

**Marie Hardt:** Methodology, Software, Formal Analysis, Writing - Original Draft, Writing - Review & Editing. **Guillermo Basulto-Elias:** Methodology, Writing - Review & Editing. **Heike Hofmann:** Methodology, Software, Visualization, Writing - Original Draft, Writing - Review & Editing. **Shauna Hallmark:** Writing - Original Draft, Writing - Review & Editing, Project Administration. **Anuj Sharma:** Supervision. **Jeffrey D. Dawson:** Validation, Writing - Review & Editing. **Matthew Rizzo:** Conceptualization, Writing - Review & Editing, Funding Acquisition. **Jun-Ha Chang:** Writing - Review & Editing, Project Administration.

## Consent Statement

All participants provided and signed an informed consent.

## Appendix A

**Figure A.1:**
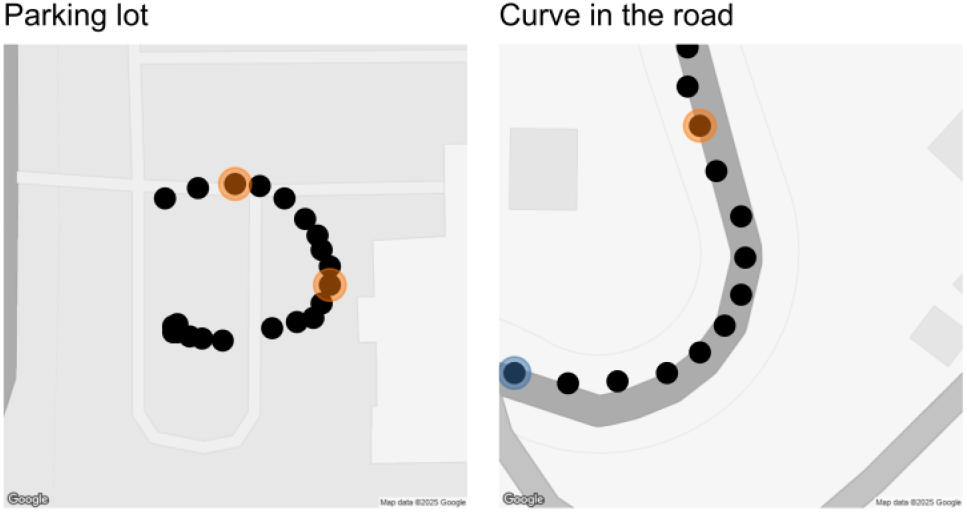
Left (blue) and right (orange) turns detected in a parking lot (left image) and on a curved road (right image).

**Table A.1:** Vertical movement of a car within 1 second under a 10^°^ change in direction.

| Speed [mph] | Offset [ft] | Offset [m] |
| --- | --- | --- |
| 10 | ± 2.5 | ± 0.8 |
| 20 | ± 5.1 | ± 1.6 |
| 30 | ± 7.6 | ± 2.3 |
| 40 | ± 10.2 | ± 3.1 |
| 50 | ± 12.7 | ± 3.9 |

NOTE. Offset values are rounded to 1 decimal place.

## Data Availability

The data used in this study are sensitive and thus cannot be shared.

